# Non-physical Intimate Partner Violence and Long-term Public Healthcare Costs in a Representative Sample of Canadian Women

**DOI:** 10.1101/2024.07.31.24311289

**Authors:** Nicholas Metheny, Gabriel John Dusing, Beverley M. Essue, Patricia O’Campo

## Abstract

This study investigated the impact of non-physical intimate partner violence (IPV), including emotional and verbal abuse, and coercive/controlling behaviors, on Ontario Health Insurance Plan costs, the universal healthcare provider in the province of Ontario, Canada. Women exposed to non-physical IPV alone had 17% higher healthcare costs over 10 years compared to those not exposed, translating to CA$686 million in additional annual costs, challenging the perception that non-physical IPV is less harmful than physical forms. We argue for prevention of non-physical IPV and improved screening in healthcare settings is vital to mitigate its long-term impacts on individuals and healthcare systems.

## Introduction

The World Health Organization (WHO) defines intimate partner violence (IPV) as “behavior by an intimate partner or ex-partner that causes physical, sexual, *or psychological harm,* including physical aggression, sexual coercion, *psychological abuse, and controlling behaviors*,”. While the WHO estimates 27% of women globally have been affected by physical and/or sexual IPV, rates of psychological harm remain largely unaccounted for in global prevalence estimates (Sardinha et al., 2022). Stemming from this lack of adequate measurement is an even more pronounced deficit in knowledge regarding the impacts of such “non-physical” forms of IPV. This gap in understanding extends to the potential public health implications, including the burden on healthcare systems and associated costs.

Physical forms of IPV (i.e., physical and sexual violence) often result in immediate bodily harm (Gujrathi et al., 2022; Nesca et al., 2021) and are shown to cause long-term, adverse physical and mental health effects (Devries et al., 2013; Gezinski et al., 2021; Kaplan et al., 2022; Nguyen et al., 2022; Rivara et al., 2019; Wuest et al., 2008). These health consequences directly lead to increased health service use, thereby driving up public healthcare costs. In some studies, such harms can be directly linked (via medical records and survey data) to IPV exposure, exposing the temporal direction of these relationships (Alonso-Borrego & Carrasco, 2023; Hisasue et al., 2024; William et al., 2022). Accurately measuring the impacts of non-physical IPV, which encompasses psychological violence, emotional abuse, stalking behaviors, coercive control, and technology-facilitated violence, is challenging in healthcare settings. Furthermore, screening for IPV in healthcare settings remains inconsistent (Coker et al., 2007; MacMillan, 2006; Perone et al., 2022). Clark et al. (2020) highlight that knowledge of how and why to screen for non-physical IPV is not commonly held among healthcare professionals, meaning that many cases go unaccounted for and lead to selection bias that blunts measurement of the true impact of these forms of IPV.

Furthermore, the variability in the types of IPV covered by healthcare screening tools (MacMillan, 2006; Rabin et al., 2009) and cultural differences in defining violence (World Health Organization, 2005) also contribute to significant underreporting of non-physical forms of IPV, and subsequently its impact on health outcomes.

Despite its exclusion from many estimates of the prevalence, incidence, and impacts of IPV, there is research that suggests non-physical IPV may be just as harmful as physical forms of violence. In one study, women who reported only psychological abuse (e.g., insults, verbal degradation, threats, restriction of autonomy) experienced similar rates of moderate to severe depression and anxiety as those exposed to both physical and psychological abuse, with both groups having worse outcomes than women reporting no abuse (Pico-Alfonso et al., 2006). Another study found women who experienced emotional abuse more than once a week had a risk of depression more than five times higher than those who experienced such abuse less frequently or not at all (Estefan et al., 2016a). In Coker and colleagues’ seminal 2000 study, women exposed to non-physical IPV alone experienced similar rates of arthritis, chronic pain, and chronic headaches as women exposed to both physical and non-physical forms of IPV (Coker, 2000). However, the long-term health effects of experiencing non-physical abuse in the absence of physical and/or sexual violence and its burden have yet to be thoroughly investigated (Stubbs & Szoeke, 2022). A better understanding of the impacts of non-physical IPV on individuals, families, and health systems may help improve efforts to properly screen for these types of IPV, refer survivors to services, and create interventions designed to prevent non-physical forms of IPV, its downstream health effects, and to mitigate the risk of later exposure to physical violence (Cascardi & Avery-Leaf, 2019; Saint-Eloi Cadely et al., 2020).

At an individual level, the psychobiology of stress model may explain the causal link between exposure to non-physical IPV and poor health outcomes, which in turn may lead to increased public healthcare costs (Kemeny, 2003; O’Connor et al., 2021). Evidence suggests that IPV acts as a chronic stressor, with measurable effects on biological markers of allostatic load (Yim & Kofman, 2019). Acting as a traumatic event, exposure to IPV has been shown to dysregulate the autonomic nervous system, releasing hormones like epinephrine, which prepare the body’s “fight-or-flight” response. Specifically, women exposed to physical, sexual, and psychological IPV show HPA axis dysregulation (HPA), marked by lower hair cortisol concentration (indicating hypocortisolism), with differences in cortisol concentration associated with the duration and intensity of IPV exposure (Alhalal & Falatah, 2020). HPA axis dysregulation, notably hypocortisolism, is observed in individuals exposed to trauma or chronic stress (Fries et al., 2005; Heim et al., 2000). Research from Cameroon (Wadji et al., 2021) and Portugal (Pinto et al., 2016) showed similar results, though none of these studies differentiated results by IPV typology. However, these responses, when left unmitigated, lead to chronic inflammation, the long-term health effects of which include cardiovascular diseases (Alfaddagh et al., 2020; Roifman et al., 2011), diabetes (Alexandraki et al., 2008), and hypertension (Xiao & Harrison, 2020). As with many conditions, access to the social determinants of health (e.g., housing and food security, social support, education, employment) (Frankenhaeuser, 1986; Kaplan et al., 2022; Kemeny, 2003), influences the severity of these health effects (Davies et al., 2015).

Using representative survey data of women in Toronto, Canada linked to longitudinal public healthcare data, this study aims to begin closing the gap in understanding regarding the harms of non-physical IPV at the community and population levels by (1) investigating the impact of non-physical IPV on the the public healthcare system in Ontario, as measured by Ontario Health Insurance Plan (OHIP) costs over a 10-year period; and (2) examining the role of neighborhood material deprivation as a marker of social determinants of health in moderating the relationship between non-physical IPV exposure and healthcare costs. Given the evidence, we hypothesize that (1) women who have experienced only non-physical IPV within 2 years of survey participation will have higher OHIP costs over the subsequent 10-year period compared to those who have not experienced any IPV, and (2) the relationship between exposure to only non-physical IPV and OHIP costs may be influenced by neighborhood material deprivation.

## Methods

### Study Sample

Our study sample (N=833) was built using participants from the Neighbourhood Effects on Health and Well-being (NEHW) study, a representative study of 50 Toronto neighborhoods conducted between 2009-2011 (O’Campo et al., 2015). Of the N=2,412 participants in NEHW, the study sample consists respondents who (1) identified as women, (2) provided a valid Ontario Health Insurance Plan (OHIP) number at the time of survey completion, and (3) provided valid responses on the IPV screener (described below). Data from the NEHW study was linked to each respondent’s OHIP record and their cumulative healthcare cost over the following ten years were calculated by IC/ES (formerly known as “Institute for Clinical Evaluative Sciences”), the organization authorized by the government of Ontario to collect and manage healthcare data for the province (IC/ES, n.d.).

### Exposure

The primary exposure of this present study was non-physical IPV (including emotional and verbal abuse as well as controlling and coercive behaviors) within two years of survey participation. It is important to note that the definition of non-physical IPV can vary by cultural and national contexts (World Health Organization, 2005). For this study, we adopt the definition outlined by the Partner Abuse Scale, details of which are provided below (Attala et al., 1994).

In the NEHW study, IPV was assessed in two stages following the example of Kirst and colleagues (Kirst et al., 2015). First, IPV within the last 10-years was assessed using a modified version of the short-form Hurt, Insult, Threaten, Scream (HITS) screener (Shakil et al., 2005; Sherin et al., 1998) that included an additional question about the restriction of actions by a partner (Yakubovich et al., 2021). If IPV was present, respondents were then asked if the exposure occurred within the last two years. If so, physical and non-physical IPV were assessed through an abbreviated version of the Partner Abuse Scale (Attala et al., 1994). The Partner Abuse Scale: Non-physical (PASNP) sub-scale measures the frequency of experiences with non-physical partner abuse, including emotional abuse, coercion, control, and jealousy. Some examples of questions from the scale include being asked if their partner: “*Feels that I should not work or go to school*,” “*Screams or yells at me*,” and “*Insults or shames me in front of others*.” In total, participants rated thirteen such behaviors on a 7-point Likert scale that indicated the frequency of each behavior. Following guidelines established by Attala et al. (1994), this score was then dichotomized – wherein a PASNP score above 15 was set as the cutoff as a compromise between maintaining high sensitivity (98.9%) and specificity (88%) while keeping the rate false positives and negatives low (12.0% and 2.2%, respectively). In our study, women reporting non-physical IPV (and no other form) within 2-years of participating in the study were considered the exposure group, and women who reported no IPV based on the HITS screener were considered the comparison group. For context, the 2-year prevalence rate of exposure to any non-physical IPV among women participating in NEHW was 20.64%. This rate includes women who experienced non-physical IPV alone as well as the n=57 women who experienced both non-physical and physical IPV excluded from our analysis.

### Outcome

The main outcome of this study was the difference in public healthcare expenditure associated with non-physical IPV exposure, assessed through the natural logarithm of the cumulative OHIP cost (standardized to 2019) for each participant from the time of participation in NEHW to the end of the study period (March 2020). The natural logarithm was chosen because it facilitates interpretation of coefficients in terms of percentage increases (Gelman et al., 2021, p. 191). For example, a coefficient of 0.10 would indicate approximately 10% higher healthcare costs for those exposed to non-physical IPV compared to those not exposed. This approach may allow better comparability with other studies than absolute dollar differences. OHIP is the universal healthcare provider for the province of Ontario, covering a wide variety of medical expenses, including visits to hospitals, emergency departments, and community health providers (Government of Ontario, n.d.). OHIP includes nearly all residents of Ontario aged 18 and older, including those who are Canadian citizens and permanent residents, as well as refugees and non-Canadians with employment authorization and their dependents. Importantly, OHIP does not include coverage for pharmaceuticals, dental care, and mental health services (with limited exceptions).

### Covariates

Models included adjustment for the following economic and socio-demographic characteristics at the time of survey completion: age, marital status, foreign-born status, employment status, estimated annual household income, and number of children. Regression models adjusted for the duration of OHIP eligibility to account for potentially lower public costs in individuals with shorter eligibility, such as those who recently relocated to Ontario or individuals whose immigration status may be tied to short-term employment. Regression models included adjustment for the Charlson Comorbidity Index as a potential confounder (Charlson et al., 2014, 2022; Sundararajan et al., 2004). Comorbid conditions have been shown to be associated with increased healthcare burden (Charlson et al., 2014; Sundararajan et al., 2004).

### Neighborhood Material Deprivation

We hypothesized that neighborhood-level material deprivation influences the relationship between non-physical IPV exposure and OHIP costs. This is supported by empirical evidence showing that individuals’ healthcare utilization (Hatef et al., 2023; Zhang et al., 2020) and IPV risk (Bonomi et al., 2014; Kirst et al., 2015; Yakubovich et al., 2020) vary with neighborhood deprivation levels. In this study, neighborhood-level material deprivation was measured by the Ontario Marginalization Index (ON-Marg), constructed using data from sources including the census and tax filings (Matheson, 2011). ON-Marg has been utilized in various studies linking neighborhood-level marginalization to health outcomes (Miao et al., 2023; Rotenberg et al., 2022; Zygmunt et al., 2020), and encompasses variables that include the proportion of adults who did not complete secondary education, proportion of single-parent families, government income assistance, unemployment, low-income status, and housing in need of major repairs. ON-Marg data was collected at the dissemination area level. Dissemination areas represent the smallest distinct units collected by the Canadian census (Government of Canada, 2021), each with approximately 400-700 residents, and serve as proxies for neighborhoods. The material deprivation indicator classifies areas into quintiles from least (first quintile) to most (fifth quintile) materially deprived.

### Statistical Analyses

For all analyses in this study, alpha was set at 5%. Descriptive statistics compared characteristics between two groups: those reporting no IPV and those reporting non-physical IPV within 2 years of NEHW survey completion, and the unadjusted association between each characteristic and exposure was evaluated through the chi-square test for categorical variables and the Kruskall-Wallis test for continuous variables.

We examined the adjusted relationship between exposure to non-physical IPV within 2-years of survey completion on individual OHIP costs over time using mixed-effects linear regression. This approach not only considers individual-level characteristics but also addresses the experiences of individuals grouped in neighborhoods with similar patterns of material deprivation. Specifically, the model allows for the estimation of distinct slopes and intercepts within each deprivation quintile, thus capturing potential variations in OHIP costs across neighborhoods having similar profiles of material deprivation.

### Sensitivity Analyses

First, considering the potential influence of neighborhood-level access to material resources on both IPV exposure and healthcare utilization, we conducted linear regression analyses stratified by quintiles of material deprivation in ON-marg. Next, given the inextricable link between comorbidities and healthcare utilization, we repeated our main analyses on the subpopulation restricted to those with no comorbidities (at the time of participation in NEHW) and, separately, those with the most (three or more).

## Results

### Descriptive Statistics

Table 1 presents the characteristics of our study sample by exposure. Among the 833 women in our study sample, 124 (14.89%) reported exposure to only non-physical IPV within 2-years of participating in the survey. This rate is lower than the overall 2-year prevalence of any non-physical IPV (20.64%), which includes the 57 women exposed to both non-physical and physical IPV who were excluded from our study sample. Significant factors associated with exposure included age, marital status, immigrant status, number of children, and household income. The exposed group appeared to be younger on average (mean: 47.91; standard deviation, SD: 10.15) compared to the comparison group (mean: 50.29; SD: 10.42) (p-value, p=0.016). The exposed group had a higher proportion of married women relative to the comparison group – 84.68% compared to 59.66% (p<0.001). There was a lower proportion of foreign-born women in the exposed group (24.19%) compared to the comparison (37.94%) (p=0.003). More (62.09%) women in the exposed group had at least 2 children compared to 46.68% of women in the comparison group (p=0.007). Women in the exposure group also appeared to be wealthier, with 71.77% reporting an annual household income of at least $74,000 as opposed to 56.84% of the comparison group (p<0.001).

**Table 1:** Sample Characteristics at Time of Survey Completion by IPV status.

|  | **No IPV reported at survey participation** | **Non-physical IPV Alone within 2 Years of Survey Participation** | **Total** | **p-value<sup>+</sup>** |
| --- | --- | --- | --- | --- |
| **Distribution by IPV Status**<br>Frequency (Row %) |  |  |  | NA |
|  | 709 (85.11%) | 124 (14.89%) | 833 (100.00%) |  |
| **Log of OHIP Costs**<br>Mean (Standard Deviation, SD) |  |  |  | 0.303 |
|  | 10.50 (0.97) | 10.60 (0.91) | 10.51 (0.96) |  |
| **Age**<br>Mean (SD) |  |  |  | 0.016 |
|  | 50.29 (10.42) | 47.91 (10.15) | 49.95 (10.4) |  |
| **Log OHIP Eligibility**<br>Mean (SD) |  |  |  | 0.119 |
|  | 12.38 (1.42) | 12.59 (1.04) | 12.41 (1.37) |  |
| **Marital Status**<br>Frequency (Column %) |  |  |  | p<0.001 |
| Unmarried | 286 (40.34%) | 19 (15.32%) | 305 (36.61%) |  |
| Married | 423 (59.66%) | 105 (84.68%) | 528 (63.39%) |  |
| **Foreign-born**<br>Frequency (Column %) |  |  |  | 0.003 |
| No | 440 (62.06%) | 94 (75.81%) | 534 (64.11%) |  |
| Yes | 269 (37.94%) | 30 (24.19%) | 299 (35.89%) |  |
| **Employment Status** |  |  |  | 0.542 |
| Frequency (Column %) |  |  |  |  |
| Unemployed | 215 (30.32%) | 41 (33.06%) | 256 (30.73%) |  |
| Employed | 494 (69.68%) | 83 (66.94%) | 577 (69.27%) |  |
| **Number of Children**<br>Frequency (Column %) |  |  |  | 0.007 |
| 0-1 | 378 (53.31%) | 47 (37.9%) | 425 (51.02%) |  |
| 2 | 215 (30.32%) | 50 (40.32%) | 265 (31.81%) |  |
| 3+ | 116 (16.36%) | 27 (21.77%) | 143 (17.17%) |  |
| **Charlson Comorbidity Index**<br>Frequency (Column %) |  |  |  | 0.132 |
| None | 131 (18.48%) | 31 (25.00%) | 162 (19.45%) |  |
| 1-2 | 350 (49.37%) | 62 (50.00%) | 412 (49.46%) |  |
| 3+ | 228 (32.16%) | 31 (25.00%) | 259 (31.09%) |  |
| **Household Income**<br>Frequency (Column %) |  |  |  | 0.002 |
| Under \$74,000 | 306 (43.16%) | 35 (28.23%) | 341 (40.94%) | |
| At Least \$74,000 | 403 (56.84%) | 89 (71.77%) | 492 (59.06%) | |
| **Neighborhood-level Material Deprivation Quintiles<sup>^</sup>**<br>Frequency (Column %) |  |  |  | 0.254 |
| 1 (Least Materially-Deprived Neighborhoods) | 159 (22.43%) | 20 (16.13%) | 179 (21.49%) |  |
| 2 | 130 (18.34%) | 19 (15.32%) | 149 (17.89%) |  |
| 3 | 105 (14.81%) | 17 (13.71%) | 122 (14.65%) |  |
| 4 | 147 (20.73%) | 34 (27.42%) | 181 (21.73%) |  |
| 5 (Most Materially-Deprived Neighborhoods) | 168 (23.70%) | 34 (27.42%) | 202 (24.25%) |  |

<sup>+</sup> p-values derived from chi-square tests for categorical variables and Kruskal-Wallis for means to assess the unadjusted differences in the distribution of characteristics between those exposed to non-physical IPV alone within 2 years of participation in NEHW and those who did not meet the threshold for IPV exposure on the HITS screener.
<sup>^</sup> Based on quintiles provided in the 2011 Ontario Marginalization Index (Matheson, 2011).

### Model Results

Table 2 presents estimates for mixed-effects regression, respectively separating the results for fixed- and random-effects. Those who reported exposure to only non-physical IPV had OHIP costs that were approximately 17% higher in the subsequent 10-years compared to those who reported no IPV within 2 years of survey completion (lJ=0.167; 95%CI: 0.064-0.270). Low intra-class correlation (ICC: 3.23E-16) suggests that variations in OHIP costs are predominantly within deprivation quintiles, rather than between them.

**Table 2:**
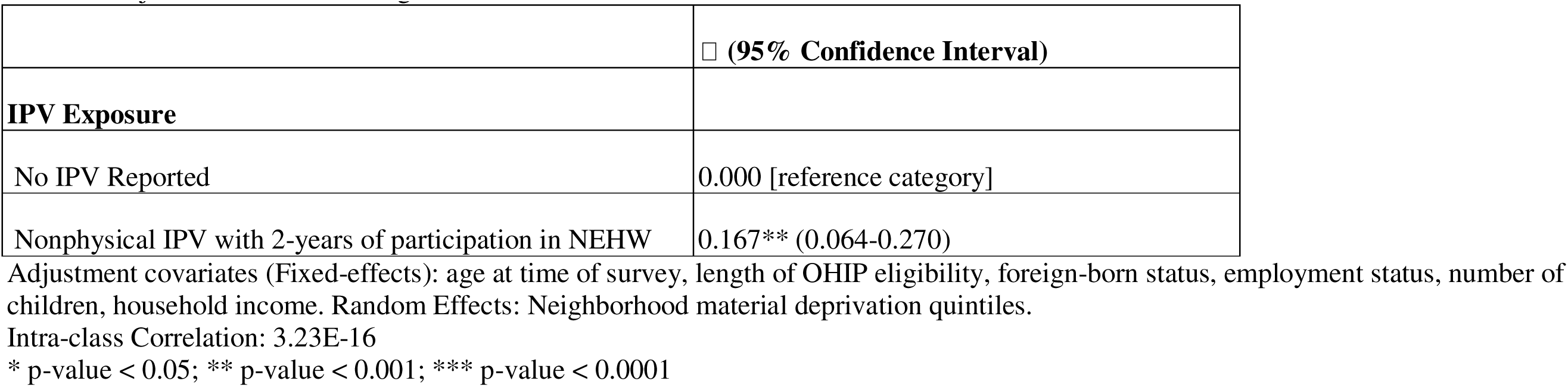
Adjusted mixed-effects regression results.

### Sensitivity Analyses

Supplementary Tables 1-5 show the results of subgroup analyses by neighborhood material deprivation quintiles. Linear regression models were estimated for each quintile separately. Positive associations between non-physical IPV exposure and healthcare costs were found across all quintiles, with point estimates ranging from 0.004 to 0.268. However, these associations did not reach statistical significance in any of the quintiles.

Lastly, Supplementary Tables 6 and 7 present the results of subgroup analyses by Charlson comorbidity index. Linear regression models were fit for women with no comorbidities (index of zero) and those with a high comorbidity burden (index of three or higher). A non-significant positive association was observed for women with no comorbidities (lJ=0.133; 95% CI: -0.115 to 0.380). Similarly, women with a high comorbidity burden, a stronger positive association was found (lJ=0.373; 95% CI: 0.018-0.728).

The sensitivity analyses demonstrate the consistency of the positive association between non-physical IPV exposure and healthcare costs across various subgroups, albeit with varying magnitudes. While the main IPWRA analysis confirmed the significant positive association, the subgroup analyses suggest potential heterogeneity in the strength of the association across levels of neighborhood material deprivation and comorbidity burden. However, it is important to note that the associations in the subgroup analyses did not reach statistical significance, likely due to the loss of statistical power.

## Discussion

Non-physical IPV (as indicated by the PASNP, which includes emotional abuse, verbal abuse, and controlling behaviors) constitutes some of the most common forms of violence (Carney & Barner, 2012; Tutty et al., 2023) and our findings indicate that it incurs excess costs on public health systems, even in the absence of reported physical and sexual violence. This is an important finding because non-physical IPV is commonly thought (by health care providers and by survivors of such violence) to be less “severe” than physical or sexual violence, causing delays in help-seeking and care for women exposed to non-physical violence (Duterte et al., 2008; García-Díaz et al., 2017; Prosman et al., 2014). This study is among the first to suggest that non-physical IPV, in the absence of physical and sexual IPV, has real and measurable impacts at a population level. This work builds on findings from previous qualitative (Halliwell et al., 2021; Tutty et al., 2023) and quantitative (Estefan et al., 2016b; Spencer et al., 2024; Tutty et al., 2023) studies suggesting that non-physical IPV has significant impacts on the health and well-being of individual women over time. We extend this work by finding that public healthcare expenditures are nearly 20% higher for women exposed to non-physical IPV with similar health profiles. This underscores the importance of prioritizing investments to address non-physical forms of violence as part of a comprehensive systems response to IPV that includes a whole-of-government effort to reduce gender inequality as a fundamental cause of IPV as well as important mediators such as poverty, educational attainment, and strong social welfare (UN Women, 2023).

Based on data provided by the Canadian Institute for Health Information, Ontario’s mean annual public health expenditure per person over the study period, adjusting for age and sex, was CA$4,086 (Canadian Institute for Health Information, 2021). Hence, the 16.7% increase in OHIP costs found in our study translates to approximately CA$654 in additional annual spending per woman. Ontario’s female population has remained at approximately 7 million since 2011 (Statistics Canada, 2017), and assuming 15% of women experience only non-physical IPV within the past two years (as in our study), we estimate exposure to non-physical IPV alone adds CA$686 million annually to OHIP’s costs, comparable to the CA$651 million the Ontario government allocated toward all hospitals and home/community care province-wide in 2019 (Government of Ontario, 2019). Extending this across the study period, this excess cost amounted to CA$9.2 billion, indicating a substantial burden of non-physical IPV exposure on Ontario’s public healthcare provider.

The prevalence of experiencing non-physical IPV without its physical forms in our study (15%) is similar to estimates from studies in other settings (Coker, 2000, p. 20; Coker et al., 2002). Experiencing non-physical IPV alone can be predictive of later exposure to both non-physical and physical forms of IPV. Prior studies indicate that past non-physical IPV is predictive of future exposure its physical forms, indicating escalation of abusive behaviors over time (Cascardi & Avery-Leaf, 2019; Saint-Eloi Cadely et al., 2020). In our descriptive analyses, women with multiple children, women with lower household incomes, and those born outside Canada reported non-physical IPV more often, lending credence to the stress-response paradigm of IPV, wherein lack of access to the social determinants of health is associated with reporting non-physical IPV (Davies et al., 2015). Interestingly, chronic disease status and neighborhood marginalization status were not significantly associated with non-physical IPV. This contrasts with previous research indicating associations between neighborhood-level structural factors, like access to material resources (Bonomi et al., 2014; Cunradi et al., 2000; Yakubovich et al., 2020), and the presence of chronic conditions and comorbidities (Gerino et al., 2018; Hartwell et al., 2023) with differential IPV risk. We posit that these discrepancies may stem from the inclusion of both physical and sexual IPV in these studies, while ours focuses solely on non-physical IPV. It may be that non-physical IPV is more equally distributed across marginalization quintiles than physical and sexual IPV. It is also possible that this more holistic measurement of marginalization is not directly comparable to the more discrete instruments used in previous studies. As for factors associated with increased healthcare costs over time, those who were unemployed at the time of the NEHW study, as well as those with more chronic conditions, had significantly higher healthcare costs than those who were employed and had fewer chronic conditions (respectively). While these were unsurprising given the extant literature, the multidimensional factors influencing non-physical IPV and related impacts on public healthcare costs means that targeted non-physical IPV mitigation strategies for couples dealing with chronic multimorbidity and precarious employment may be warranted.

Investing in interventions that address non-physical IPV at both individual and systemic levels, targeting factors such as social norms, economic inequalities, and institutional practices, could result in cost savings for the public healthcare system, and subsequently, the Canadian taxpayer by interrupting cycles of violence. First, these could include provisions for dyadic interventions that have been shown to rehabilitate dysfunctional relationships (Hayes et al., 2015; Levitov & Fall, 2020) and promote healthy relationship dynamics (Di Maio et al., 2024; Monk et al., 2018), thus, preventing future incidences of non-physical IPV and the escalation of non-physical IPV to physical and or sexual violence (O’Leary, 2015). Furthermore, risk factors for male perpetration of non-physical IPV toward female partners include exposure to violence in childhood, post-traumatic stress disorders, personality disorders, substance use, and patriarchal attitudes toward gender norms (Clare et al., 2021; Fulu et al., 2013; Spencer et al., 2024). Investment in programs that address these factors could also serve as a means to prevent non-physical IPV. Examples of such programs have shown promise in low- and middle-income settings (Doyle & Kato-Wallace, 2021) and could be adapted for high-income settings such as Canada.

Second, several systematic reviews highlight an important gap with regard to routine screening and referral for a non-physical IPV (Sprague et al., 2012; Waalen et al., 2000) in clinical settings. Structural interventions could include expanding healthcare education and in-service training to train health professionals on the warning signs of non-physical forms of IPV to facilitate early identification, referrals, and support (Arkins et al., 2016). Considering non-physical IPV often portends the co-occurrence with and/or escalation of IPV to physical and sexual forms (Cascardi & Avery-Leaf, 2019; Saint-Eloi Cadely et al., 2020), the availability of services for the psychological and economic empowerment of those exposed to non-physical IPV are also needed. This may help women repair or exit violent relationships earlier, reducing the long-term impacts of IPV exposure. At a societal level, strategies to support equitable access to the social determinants of health, especially employment–which facilitates access to employer-provided private insurance for services not covered by OHIP– can help women in violent relationships of all kinds empower themselves to assess their options and choose what is best for them and their families (Eggers del Campo & Steinert, 2022).

There are several limitations to the current study. First, non-physical IPV varies widely in definition and can be context- and culture-specific (Doolabh et al., 2022). The lack of a common, comprehensive definition of non-physical IPV means that forms of non-physical IPV are likely missing from the PASNP, and therefore from this analysis. One important example includes online forms of non-physical IPV, which have become increasingly prevalent in recent years (Damra et al., 2023). Second, exposure to non-physical IPV was assessed only at the time of participation in the NEHW study, and hence, it is important to acknowledge that subsequent exposures would be missed due to limitations inherent in the study design. Consequently, we cannot rule out the presence of non-physical IPV among the comparison group during the 10 years after the survey was completed, which suggests that the impacts measured are likely underestimated. While International Classification of Disease (ICD) codes exist for physical and sexual violence (Olive, 2018), they are infrequently used in medical charts and no such codes exist for non-physical IPV. This precludes our ability to detect exposure to non-physical IPV over time or to understand how exposure to non-physical IPV in NEHW might lead to physical or sexual IPV in the future. Longitudinal studies of survivors of non-physical IPV are needed to elucidate a more precise health and economic impact over time as well as trajectories of IPV exposure. Third, individuals may have been incorrectly assigned to the comparison group due to nondisclosure, in line with historical underreporting of IPV (Agüero & Frisancho, n.d.; Chan, 2011), which is also a limitation acknowledged in the validation of the PASNP (Attala et al., 1994). Together, these limitations likely mean the findings reported here are underestimates of their true impacts. However, the long-term follow-up, robust methods, and population-based sample used in this analysis strengthen our conclusions and recommendations, and serve as a foundation on which future research, advocacy, and intervention development can be built. Furthermore, the consistency of the positive association found in our main analysis and subgroup analyses is a strength of our study. Additionally, although non-significant due to the likely loss of statistical power, our supplementary analyses stratified by neighborhood deprivation quintiles and comorbidity levels suggest that the association between IPV and healthcare costs may be robust across different subpopulations.

## Conclusion

The majority of violence research excludes exposure to non-physical IPV, a reflection of the perceived hierarchy of severity in IPV typology. Although the effects of physical and sexual IPV are often more visible (since they are more likely to result in physical injuries), it is critical to recognize from our findings that the impacts of non-physical IPV are not less severe. Our findings demonstrate that exposure to non-physical IPV alone incurs significantly higher 10-year OHIP costs, challenging the notion that these less visible forms of violence are less harmful. Indeed, the pervasive nature of non-physical IPV— potentially due to being a less risky strategy for the perpetrator—underscores its substantial long-term impacts on health and healthcare systems. Non-physical forms of violence remain under-researched and less well-understood than physical and sexual IPV. Additional inquiry into the forms of non-physical IPV that are most prescient in specific communities could lead to additional, culturally specific quantitative measures that more accurately capture its harms within and between diverse populations, which is especially relevant in Toronto, where nearly half of residents are foreign-born (Government of Canada, 2022). Such measures should then be used to identify points of individual and dyadic intervention for the prevention and mitigation of non-physical IPV. Finally, and importantly, while our research highlights the financial costs non-physical IPV levies on healthcare systems, one must not lose sight that the profound personal cost experienced by victims and their families, including children who witness such violence, are of paramount concern and require dedicated attention and intervention.

## Statements and Declarations

### Consent to participate

All participants provided written informed consent when they participated in the NEHW study.

### Ethical Considerations

Ethics approval was granted by the Research Ethics Board of St. Michael’s Hospital, Toronto (REB# 20-120).

## Consent for publication

Not Applicable.

## Data Availability Statement

All data produced in the present study are not publicly available due to confidentiality requirements. The data are securely housed with ICES, with access restricted by confidentiality agreements.

## Data Availability

All data produced in the present study are not publicly available due to confidentiality requirements. The data are securely housed at ICES, with access restricted by confidentiality agreements.

## Appendix: Supplementary Materials

For all models in this section, adjustment covariates are: age at time of survey, length of OHIP eligibility, foreign-born status, employment status, number of children, household income, Charlson comorbidity index.

Legend: * p-value < 0.05; ** p-value < 0.001; *** p-value < 0.0001

**Supplementary Table 1:** Adjusted linear regression results for the effect of exposure to non-physical IPV within 2 years of participation in NEHW. Subpopulation consists of those residing in neighborhoods with Quintile 1 of neighborhood-level material deprivation (lowest material deprivation)

|  |  |
| --- | --- |
|  | □<br>(95% Confidence Interval) |
| **IPV Exposure** |  |
| No IPV Reported | 0.000 [reference category] |
| Nonphysical IPV within 2-years of participation in NEHW | 0.268<br>(-0.179, 0.716) |

**Supplementary Table 2:** Adjusted linear regression results for the effect of exposure to non-physical IPV within 2 years of participation in NEHW. Subpopulation consists of those residing in the second-to-least materially deprived neighborhoods (Material Deprivation Quintile = 2)

|  |  |
| --- | --- |
|  | □<br>(95% Confidence Interval) |
| **IPV Exposure** |  |
| No IPV Reported | 0.000 [reference category] |
| Nonphysical IPV within 2-years of participation in NEHW | 0.148<br>(-0.294, 0.589) |

**Supplementary Table 3:** Adjusted linear regression results for the effect of exposure to non-physical IPV within 2 years of participation in NEHW. Subpopulation consisting of those residing in the middle-20% of materially deprived neighborhoods (Material Deprivation Quintile = 3)

|  |  |
| --- | --- |
|  | □<br>(95% Confidence Interval) |
| **IPV Exposure** |  |
| No IPV Reported | 0.000 [reference category] |
| Nonphysical IPV within 2-years of participation in NEHW | 0.249<br>(-0.113, 0.611) |

**Supplementary Table 4:** Adjusted linear regression results for the effect of exposure to non-physical IPV within 2 years of participation in NEHW. Subpopulation consisting of those residing in the fourth-most materially deprived neighborhoods (Material Deprivation Quintile = 4)

|  |  |
| --- | --- |
|  | □<br>(95% Confidence Interval) |
| **IPV Exposure** |  |
| No IPV Reported | 0.000 [reference category] |
| Nonphysical IPV within 2-years of participation in NEHW | 0.004<br>(-0.278, 0.286) |

**Supplementary Table 5:** Adjusted linear regression results for the effect of exposure to non-physical IPV within 2 years of participation in NEHW. Subpopulation consisting of those residing in the most materially-deprived neighborhoods (Material Deprivation Quintile = 5)

|  |  |
| --- | --- |
|  | □<br>(95% Confidence Interval) |
| **IPV Exposure** |  |
| No IPV Reported | 0.000 [reference category] |
| Nonphysical IPV within 2-years of participation in NEHW | 0.218<br>(-0.138, 0.573) |

**Supplementary Table 6:** Adjusted linear regression results for the effect of exposure to non-physical IPV within 2 years of participation in NEHW. Subpopulation: Women with a Charlson comorbidity index of zero at the time of participation in NEHW.

|  | □<br>(95% Confidence Interval) |
| --- | --- |
| **IPV Exposure** |  |
| No IPV Reported | 0.000 [reference category] |
| Nonphysical IPV within 2-years of participation in NEHW | 0.133<br>(-0.115, 0.380) |

**Supplementary Table 7:** Adjusted linear regression results for the effect of exposure to non-physical IPV within 2 years of participation in NEHW. Subpopulation: Women with a Charlson comorbidity index of three or higher at the time of participation in NEHW (Top 20% of women).

|  | □<br>(95% Confidence Interval) |
| --- | --- |
| **IPV Exposure** |  |
| No IPV Reported | 0.000 [reference category] |
| Nonphysical IPV within 2-years of participation in NEHW | 0.373*<br>(0.018, 0.728) |

